# Analysis of external quality assessment samples revealed crucial performance differences between commercial RT-PCR assays for SARS-CoV-2 detection when taking extraction methods and real-time-PCR instruments into account

**DOI:** 10.1101/2020.09.18.20185744

**Authors:** Monika Malecki, Jessica Luesebrink, Andreas Wendel, Frauke Mattner

## Abstract

In limelight of the ongoing pandemic SARS-CoV-2 testing is critical for the diagnosis of infected patients, contact-tracing and mitigating the transmission. Diagnostic laboratories are expected to provide appropriate testing with maximum accuracy. Real-time reverse transcriptase PCR (RT-PCR) is the diagnostic standard yet many commercial diagnostic kits have become available. However, only a handful of studies have reviewed their performance in clinical settings. The aim of this study was to compare the performance of the overall analytical matrix including the extraction kit (BD MAX, Promega, Qiagen), the PCR instrument (Agilent Mx3005P, BD MAX, Qiagen Rotor-Gene, Roche Cobas z 480) and the RT-PCR assay (Altona Diagnostics, CerTest Biotec, R-Biopharm AG) using predefined samples from proficiency testing organizers.

The greatest difference of the Ct values between the matrices was 9 cycles. One borderline sample could not be detected by 3 out of 12 analytical matrices and yielded a false negative result. We therefore conclude that diagnostic laboratories should take the complete analytical matrix in addition to the performance values published by the manufacturer for a respective RT-PCR kit into account. With limited resources laboratories have to validate a wide range of kits to determine appropriate analytical matrices for detecting SARS-CoV-2 reliably. The interpretation of clinical results has to be adapted accordingly.

## Background

Ever since the severe acute respiratory syndrome coronavirus 2 (SARS-CoV-2) was identified as the causative agent of the coronavirus disease 2019 (COVID-19), the number of commercial kits detecting the virus from clinical samples keeps growing (1, 2). At the same time expansion of SARS-CoV-2 diagnostic testing became the top public health priority to mitigate the spread of the disease (3). Therefore laboratories have to evaluate different diagnostic assays and establish them in laboratory workflows at once. To complicate this assessment even further, laboratories were, and still are, confronted with supply shortages of disposables, technical equipment and/or reagents. These bottlenecks have led to a considerable obstacle in establishing successful routine SARS-CoV-2 diagnostic. The situation at hand requires agile management and implementation of available components, which might not yet have been validated in a clinical setting beforehand.

Accurate diagnosis and quality assurance measures are of utmost importance. External quality assessments (EQA) provided by proficiency testing organizers, designed to assess the ability of laboratories to detect a pathogenic agent at a clinically relevant level, are particularly suitable to meet these requirements. Additionally, standardized EQA samples enable a laboratory to evaluate individual components of a test without relying on the performance data of the manufacturer solely.

RT-PCR (reverse transcriptase-polymerase chain reaction) based diagnostic is officially recommended as the gold standard method for SARS-CoV-2 detection (4). However, performance characteristics of a molecular method not only depend on the preanalytics but also on the analytical matrix available in a laboratory, in case of a RT-PCR as follows: extraction method, the reagents of the downstream application (primers, targets, polymerase) and the PCR instrument. To determine the optimal test algorithm in a respective laboratory it is inevitable to compare all combinations of the aforementioned components. Subsequently, the limitations of any such combination can be identified. The results of the comparisons provide evidence for a risk assessment in case the optimal setup is not available and a less accurate combination has to be used.

In this study, two diagnostic laboratories of a tertiary care hospital equipped with different PCR systems validated the available kits on the respective systems for SARS-CoV-2 testing.

## Objectives

In order to evaluate the performance of analytical components used for SARS-CoV-2 diagnostic we compared three commercially available RT-PCR kits, six extraction kits and four PCR cyclers in all possible combinations using predefined EQA samples.

## Study design

EQA samples were received from two proficiency testing providers: the German INSTAND e.V. and Quality Control for Molecular Diagnostics (QCMD) based in the UK. The EQA samples were derived from in vitro systems, no human clinical samples were used. The distributed samples were SARS-CoV-2 positive, positive for a different human coronavirus or coronavirus negative. Viral RNA was isolated using automated Maxwell® Instruments (Promega), extracted manually with QIAamp® Viral RNA Mini Kit (Qiagen) or processed in Becton Dickinson’s BD MAX™ system. In the latter method extraction and thermocycling was performed in one instrument (Table 1). Real-time PCR instruments in use for the SARS-CoV-2 diagnostic included Rotor-Gene® Q (Qiagen), Cobas® z 480 (Roche) and Mx3005P (Agilent).

**Table 1:**
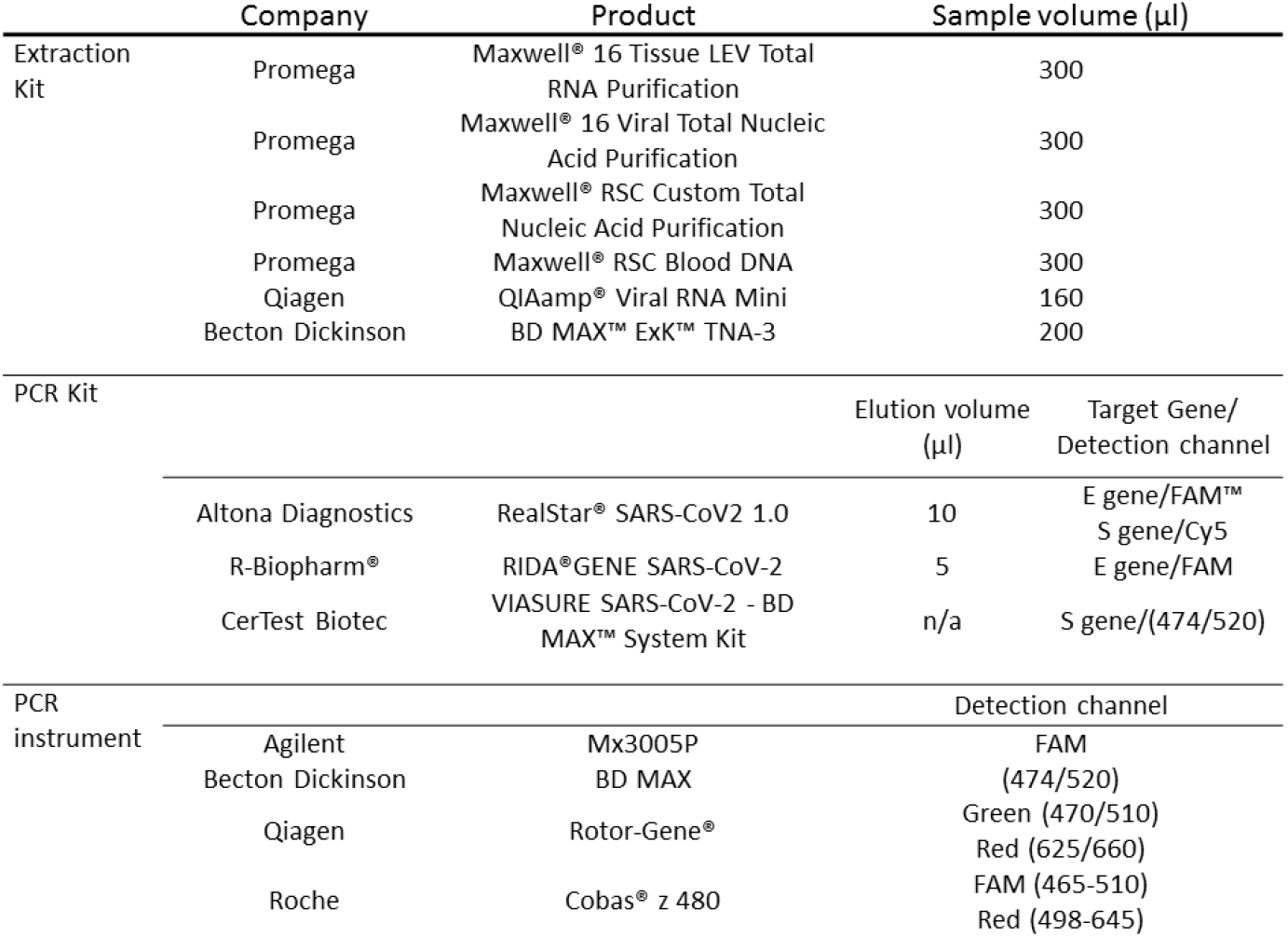
Details of the compared PCR instruments, extraction and PCR kits.

|  | Company | Product | Sample volume (µl) |  |
| --- | --- | --- | --- | --- |
| Extraction Kit | Promega | Maxwell® 16 Tissue LEV Total RNA Purification | 300 |  |
|  | Promega | Maxwell® 16 Viral Total Nucleic Acid Purification | 300 |  |
|  | Promega | Maxwell® RSC Custom Total Nucleic Acid Purification | 300 |  |
|  | Promega | Maxwell® RSC Blood DNA | 300 |  |
|  | Qiagen | QIAamp® Viral RNA Mini | 160 |  |
|  | Becton Dickinson | BD MAX™ ExK™ TNA-3 | 200 |  |
| PCR Kit |  |  | Elution volume (µl) | Target Gene/ Detection channel |
|  | Altona Diagnostics | RealStar® SARS-CoV2 1.0 | 10 | E gene/FAM™<br>S gene/Cy5 |
|  | R-Biopharm® | RIDA®GENE SARS-CoV-2 | 5 | E gene/FAM |
|  | CerTest Biotec | VIASURE SARS-CoV-2 - BD MAX™ System Kit | n/a | S gene/(474/520) |
| PCR instrument |  |  | Detection channel |  |
|  | Agilent | Mx3005P | FAM |  |
|  | Becton Dickinson | BD MAX | (474/520) |  |
|  | Qiagen | Rotor-Gene® | Green (470/510)<br>Red (625/660) |  |
|  | Roche | Cobas® z 480 | FAM (465-510)<br>Red (498-645) |  |

Three RT PCR kits were tested. The RealStar® SARS-CoV-2 RT-PCR Kit (Altona Diagnostics) follows the two-target strategy by detecting the S and E genes (encoding the spike and the envelope protein of SARS-CoV-2 respectively), whereas the RIDA®GENE SARS-CoV-2 Kit (R-Biopharm®) detects the E gene and VIASURE SARS-CoV-2 - BD MAX™ System Kit (CerTest Biotec) the S gene (Tab. 1). The extraction and RT-PCR kits were applied according to the manufacturer’s specifications. To compare the analytical matrices with each other, consisting of the variables extraction kit / PCR instrument / PCR kit, the cycle threshold (Ct) values of the EQA samples have been taken into account.

## Results

All negative samples or samples positive for a different human coronavirus were accurately detected as SARS-CoV-2-negative in every analytical matrix (data not shown). The EQA samples 1-4 (INSTAND) were accurately identified as SARS-CoV-2 positive in all analytical matrices. Six analytical matrices were performed to detect the E and the S gene. The maximum Ct value difference of the matrices was 9 cycles within sample 1 (Ct 18.64 for the matrix Promega 16 Viral Total/Rotor-Gene/Altona; Ct 27.66 for Promega RSC Custom/Mx3005P/Altona for the E gene; Ct 17.1 and Ct 26.42 for the S gene respectively) (Figure 1a and 1b).

**Figure 1:**
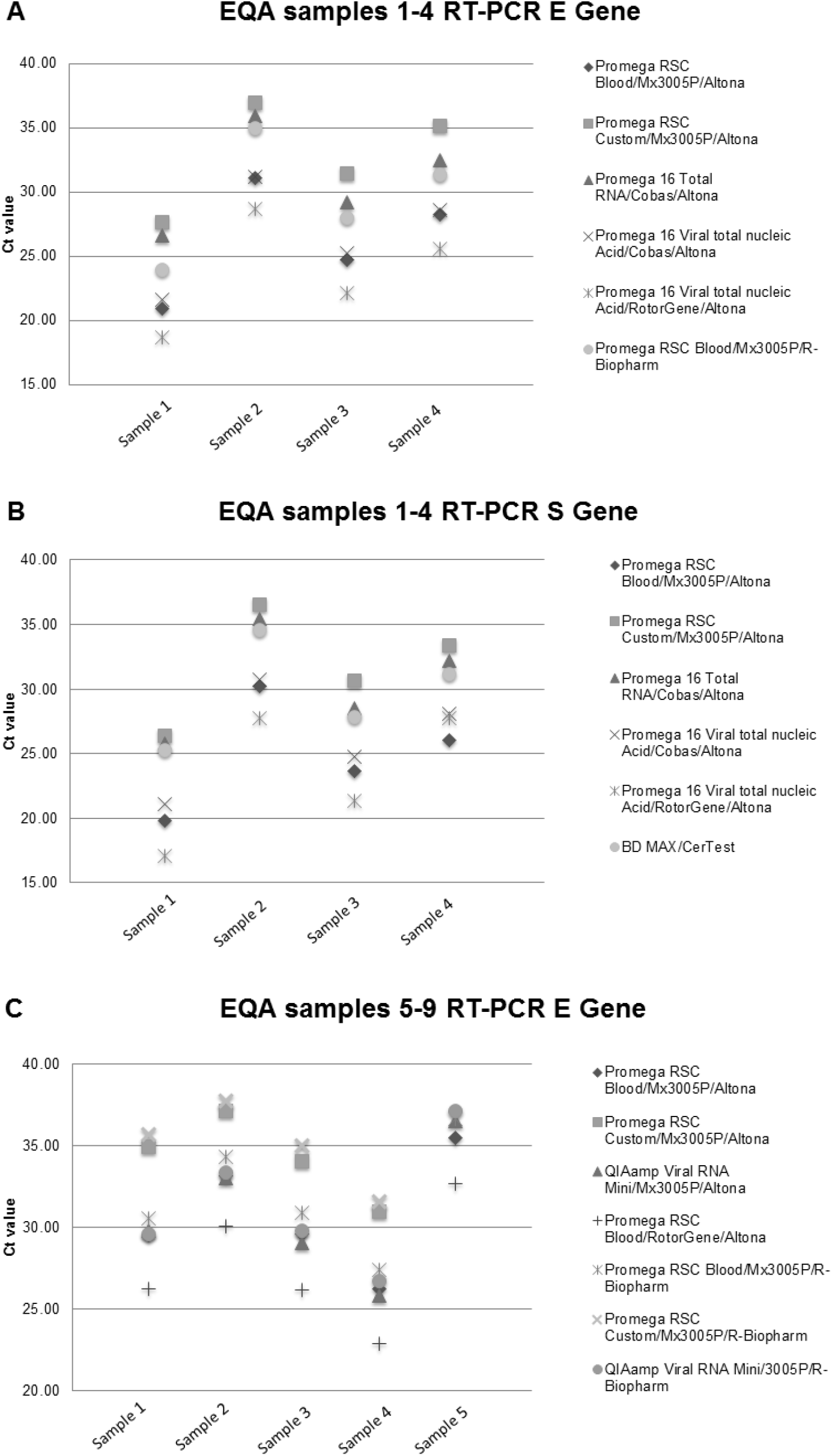
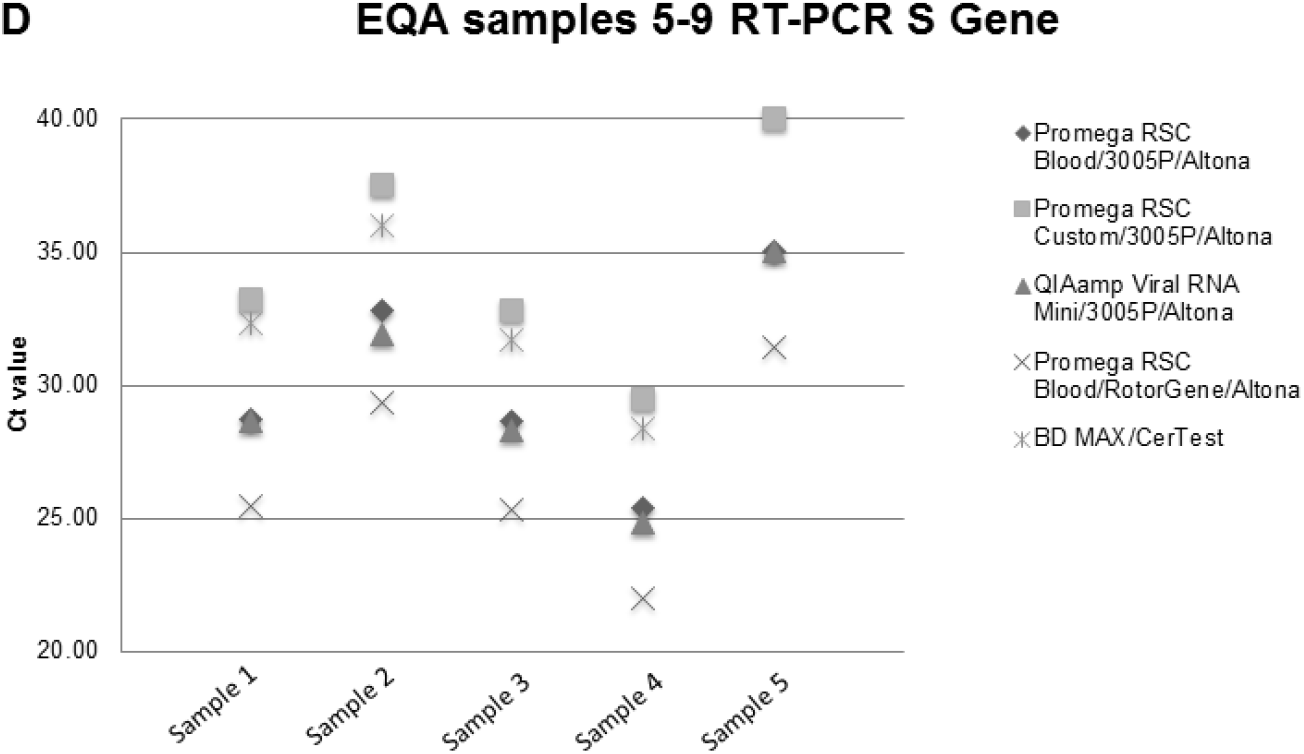
RT-PCR Kits display variations in Ct values dependent on the analytical matrix. (Legend continued on next page) EQA samples 1-4 were purchased from the proficiency testing organizer INSTAND and the samples 5-9 from QCMD. The symbols indicate the respective analytical matrix consisting of an extraction kit, the PCR instrument und the RT-PCR assay. No positive result could be obtained for sample 9 with Promega RSC Custom/Mx3005P/Altona and Promega RSC Custom/Mx3005P/R-Biopharm for the E gene and BD MAX/CerTest for the S gene (Figure 1c-d).

In the second set of samples (QCMD) 7 analytical combinations were used to detect the E gene and 5 to detect the S gene. In contrast to the aforementioned satisfactory results according to the proficiency testing provider, sample 9, defined as borderline SARS-CoV-2 positive by the organizer, was assessed as false-negative in three analytical matrices: Promega RSC Custom/Mx3005P/Altona and Promega RSC Custom/Mx3005P/R-Biopharm for the E gene and BD MAX/CerTest for the S gene (Figure 1c and 1d.). The lowest Ct value of sample 9 (31.46) was assessed for the combination of Promega RSC Blood/Rotor-Gene/Altona for the S gene (Figure 1d). According to the recommendations of the manufacturer 2 results out of 3 performed with the RIDA®GENE assay should have been assessed as inconclusive, since they showed a Ct > 35. The maximum Ct value difference of the matrices used for the QCMD samples was 9 cycles within sample 5 detecting the E gene (Ct 26.26 for the matrix Promega RSC Blood/Rotor-Gene/Altona and Ct 35.7 for Promega RSC Custom/Mx3005P/R-Biopharm) (Figure 1c).

## Discussion

Commercial RT-PCR diagnostic kits for COVID-19 have been compared before (5,6). Using SARS-CoV-2-positive clinical samples a variation of the 95 % limit of detection up to a 6-fold range between RT-PCR kits has been shown (6). In this study, unlike in other publications, the comparison also takes different extraction methods and PCR instruments into account. We were able to show a wide variation of Ct values for the three components applied respectively. The difference of up to 9 Ct values for the samples analyzed give reason for concern. Clinical samples may be reported falsely as negative, unlike indicated by the sensitivity values published in predated studies. Notably the samples with the highest Ct values or being tested as false-negative have been extracted with the Maxwell® RSC Custom Total Nucleic Acid Purification kit, which cannot be recommended for COVID-19 testing according to our findings (Figure 1). Samples extracted with the Maxwell® 16 Tissue LEV Total RNA Purification kit or processed with the BD system showed continuously high Ct values (Figure 1a, b, d), whereas the PCR kits from Altona and R-Biopharm performed on a similar level depending on the extraction method and the cycler used (Figure 1). This demonstrates that the PCR kit is not the only determinant. The extraction kits seem to influence the analytical performance of commercially available diagnostic kits considerably. Our findings also underline the necessity of EQA samples with low amounts of analytes in order to reveal less suitable methods for SARS-CoV-2 detection in routine diagnostic. Especially since Ct values are becoming increasingly relevant to assess the infectivity of SARS-CoV-2 patients and to discontinue their isolation (7). Pooling of samples for SARS-CoV-2 testing has been discussed as a strategy to overcome shortages (8). When performing pooling – a method provoking a loss of sensitivity – one should scrutinize the test algorithm even further.

The study in hand has certain limitations. Not all possible combinations of components were performed due to limited sample volumes. For the same reasons it was not possible to perform multiple testing of the respective test algorithms. It should also be noted that the volumes of the primary samples used for the extraction as well as the volumes of the eluates applied in the RT-PCR differed up to a double (Tab. 1). Lastly the performance of the test components used might have changed by the time of publication since the suppliers continuously improve these.

We therefore conclude that, despite a scarcity of resources, diagnostic laboratories have to not only implement available kits immediately and thoroughly but also to determine the effects of different extraction methods and PCR instruments in order to enhance the accuracy of the diagnostic kits in use. Moreover, the knowledge about the influence of both determinants on Ct values with regard to SARS-CoV-2 RT-PCR testing should lead to a more careful interpretation of the obtained results. During supply shortages clinical samples may be triaged depending on the clinical course of the patient. For example, therapy depending testing has to be prioritized and performed with the most accurate analytical matrix available.

## Data Availability

Data available within the article.

